# Is platelet activation a link between metabolic syndrome and cognitive impairment in patients with schizophrenia?

**DOI:** 10.1101/2023.01.10.23284409

**Authors:** Olaoluwa O. Okusaga, K. Vinod Vijayan, Rolando E. Rumbaut

## Abstract

**Introduction:** Schizophrenia is a severe psychiatric condition associated with cognitive impairment and premature dementia. Furthermore, metabolic syndrome (MetS)—combined central obesity, diabetes, dyslipidemia and hypertension—is highly prevalent in patients with schizophrenia and is believed to contribute to cognitive impairment and premature dementia in patients with schizophrenia. However, the mechanisms by which MetS contributes to cognitive impairment in patients with schizophrenia is unclear. Based on the association of MetS with platelet activation and the ability of activated platelets to impact blood-brain-barrier function, we tested the hypothesis that platelet activation is associated with both MetS and cognitive impairment in two independent pilot samples of patients with schizophrenia.

**Methods:** In the first pilot sample (sample A) we recruited 13 veterans with either schizophrenia or schizoaffective disorder with MetS (MetS+, n=6), and without MetS (MetS-, n=7). We administered the Measurement and Treatment Research to Improve Cognition in Schizophrenia (MATRICS) Consensus Cognitive Battery (MCCB) on all 13 veterans and assessed platelet activation using flow cytometry. In the second pilot sample (sample B), we identified 10 non-veteran MetS+ patients with schizophrenia and 10 age-, and sex-matched MetS-patients with schizophrenia from previously collected data on 106 patients enrolled in a non-MetS study. Participants in sample B had data on the NIH Toolbox cognitive battery (NIH Toolbox) and plasma soluble P-selectin (sP-selectin), a marker of platelet activation. We compared flow cytometry platelet activation in MetS+ and MetS- using the Mann Whitney test and the median test to compare sP-selectin and cognitive measures. We also measured the correlation between platelet activation and cognition using Spearman’s rho correlation.

**Results:** Platelet activation was significantly higher in MetS+ than MetS- (mean rank 8.60 vs. 3.83, p=0.017). Median score for the picture vocabulary test (language ability) was significantly lower in MetS+ relative to MetS- (82.35 vs. 104, p=0.015). In addition, platelet activation correlated negatively (rho = −0.74, p= 0.009) with the Wechsler Memory Scale: Spatial Span (nonverbal working memory) and plasma sP-selectin correlated negatively (rho = −0.55, p= 0.029) with the List Sorting Working Memory Test.

**Conclusion:** Our preliminary findings suggest that platelet activation is involved in the association of MetS with cognitive impairment in patients with schizophrenia. Future studies are needed to elucidate the role of platelets in MetS-related cognitive impairment in patients with schizophrenia.

## Introduction

Schizophrenia is a severe psychiatric disorder associated with cognitive impairment [1] and high prevalence of dementia [2]. In fact, there is generally a 2.5 fold increased risk of dementia [2] and at age 66 years, a 21.5 fold higher prevalence of dementia in patients with schizophrenia relative to people without a severe mental illness [3]. Schizophrenia is also associated with high prevalence of metabolic syndrome (a constellation of health conditions— including central obesity, diabetes, dyslipidemia and hypertension—that increase a person’s risk for cardiovascular disease complications) [4]. The prevalence of Metabolic syndrome (MetS) in patients with schizophrenia is 1.5-2.0 times higher than in the general population [5]. Importantly, MetS has been linked to cognitive impairment in patients with schizophrenia [6] and is believed to contribute to the high incidence and prevalence of dementia in patients [7]. However, the mechanisms underlying the association of MetS with cognitive impairment in schizophrenia are not well understood.

In non-psychiatric patients, MetS is associated with platelet activation [8,9]. Activated platelets express P-selectin, a transmembrane protein also expressed by endothelial cells [10]. Furthermore, activated platelets interact directly with leukocytes via P-selectin binding to P-selectin glycoprotein ligand-1 (PSGL-1) on leukocytes resulting in the facilitation of procoagulant and inflammatory pathways [10]. Also, platelet-leukocyte interaction involving P-selectin and PSGL-1 result in proteolytic shedding and release of soluble P-selectin (sP-selectin) into the circulation [11]. sP-selectin is biologically active and mice genetically modified to produce excessive amounts of plasma sP-selectin exhibit blood-brain-barrier (BBB) abnormalities and abnormal social behavior [12]. Plasma sP-selectin is also associated with stroke in humans [13]. Moreover, activated platelets can release growth factors, amines, cytokines and chemokines which can impact BBB function and result in neuroinflammation [14]. Consequently, platelet activation has been proposed as a potential link between MetS and neurodegeneration, leading to dementia [15].

Assessment of platelet activation by flow cytometry [16] and plasma sP-selectin [17] respectively, have been reported in schizophrenia but to our knowledge, the association of platelet activation with MetS and cognition in schizophrenia has not been reported. We therefore evaluated platelet activation, and cognitive assessments in two independent pilot samples of patients with schizophrenia or schizoaffective disorder with differing MetS status. We hypothesized that platelet activation will be associated with both MetS and cognitive impairment in patients with schizophrenia.

## Materials and Methods

### Study Participants

#### Pilot sample A

We enrolled 13 veterans with schizophrenia or schizoaffective disorder with MetS (MetS+, n=6), and without MetS (MetS-, n=7) in the outpatient clinic. We defined MetS using the National Cholesterol Education Program Adult Treatment Panel III (NCEP ATP III) criteria [18], i.e. three or more of the following: (1) Waist circumference >102 cm (men) or >88 cm (women), (2) systolic blood pressure >130 mmHg or diastolic blood pressure >85mmHg, (3) plasma triglycerides (TG) >150 mg/dL, high density lipoprotein (HDL) <40 mg /dL (men) and < 50 mg/dL (women), (4) fasting glucose level >110 mg/dL. We combined schizophrenia and schizoaffective disorder as both conditions are identical on key cognitive, social cognitive, and neural measures [19]. Inclusion criteria for this study were: (1) Male and female Veterans, (2) DSM-5 diagnosis of schizophrenia or schizoaffective disorder, (3) Age 18 to 60 years, (4) Negative urine pregnancy test in women of childbearing age, (5) clinically stable without medication changes for at least 3 months. Exclusion criteria were: (1) Pervasive developmental disorder, dementia, delirium, other cognitive disorders, (2) Current suicidal and homicidal ideation.

#### Pilot sample B

Using previously collected data on a sample of 106 non-veteran patients [17] with schizophrenia enrolled in a non-metabolic syndrome study (at the time of discharge from inpatient psychiatric treatment), we identified patients that met criteria for MetS (MetS+). The criteria used for MetS were the combination of overweight/obesity, plasma TG > 150 mg/dL, and HDL < 40 mg/dL). 10 patients met criteria for MetS. From the same sample of 106, we then identified patients without MetS (MetS-) matching those with MetS on age and sex (n=10). The original sample of 106 met the following inclusion criteria: (1) DSM-5 diagnosis of schizophrenia, (2) age between 18-60 years, (3) negative pregnancy test if female of childbearing age. The exclusion criteria were: (1) cognitive disorders such as pervasive developmental disorder, delirium, dementia, (2) current suicidal and homicidal ideations, (3) positive for psycho-stimulants such as cocaine, amphetamines and ecstasy on urine drug screen, (4) primary inflammatory conditions including any infection, neoplasm, autoimmune diseases, (5) currently taking non-steroidal anti-inflammatory medication, (6) current or anticipated corticosteroid use, (7) recent use of warfarin or any anticoagulant.

The diagnoses of schizophrenia or schizoaffective disorder were confirmed in both samples using the Mini International Neuropsychiatric Interview version 5 [20]. This study complied with all the relevant national regulations, institutional policies and in accordance with the tenets of the Declaration of Helsinki, and were approved by the authors’ institutional review board or equivalent committee. All the participants provided informed consent after a member of the research team had explained the procedures, including benefits and risks, and all their concerns and questions had been addressed.

### Assessment of cognition

In sample A, we evaluated cognition using the Measurement and Treatment Research to Improve Cognition in Schizophrenia (MATRICS) Consensus Cognitive Battery (MCCB) [21]. The MCCB was specifically developed to assess several cognitive domains in individuals with schizophrenia and related disorders [21]. The individual tests in the MCCB battery and the domains assessed are as follows: Brief Assessment of Cognition in Schizophrenia: Symbol-Coding, Category Fluency: Animal Naming, Trail Making Test: Part A (all assess Speed of processing); Continuous Performance Test—Identical Pairs (assesses Attention/Vigilance); Wechsler Memory Scale®—3rd Ed: Spatial Span, Letter–Number Span (assess nonverbal and verbal Working memory respectively); Hopkins Verbal Learning Test—Revised™ (assesses Verbal learning); Brief Visuospatial Memory Test—Revised (assesses Visual learning); Neuropsychological Assessment Battery®: Mazes (assessment of Reasoning and problem solving); Mayer-Salovey-Caruso Emotional Intelligence Test™: Managing Emotions (assesses Social cognition). T scores (available from The MCCB Scoring Program) were derived for each cognitive test. Neurocognitive composite and total MCCB composite scores (measures of broad intellectual ability) were also computed based on the individual tests. The same certified and experienced rater (blind to the MetS status of patients) administered the MCCB on all the participants.

In sample B, we evaluated cognition using the National Institute of Health Toolbox Cognitive Test Battery (NIH Toolbox) [22]. The NIH Toolbox is useful for assessing cognition across the lifespan (ages 3 to 85 years) [23] and includes the following tests of several cognitive domains: 1) Picture Vocabulary Test and Oral Reading Recognition Test both measure language skills; 2) List Sorting Working Memory Test measures working memory; 3) Dimensional Change Card Sort and Flanker Inhibitory Control and Attention Tests measure executive function-Inhibitory Control and Attention; 4) Pattern Comparison Processing Speed Test measures processing speed; 5) Picture Sequence Memory Test measures episodic memory. Crystallized Cognition Composite, Fluid Cognition Composite, and Total Cognition Composite scores were also computed from the Toolbox measures [24]. Crystalized cognition is related to a person’s store of knowledge and experiences and tends to be stable in old age [24]. On the other hand, fluid cognition reflects ability to register and manipulate new information to solve everyday problems and tends to be sensitive to biological processes (e.g. aging, and neurological disorders) that affect current brain function [24]. We collected fully adjusted scores on each cognitive domain for each research participant. Fully adjusted scores are computed (automatically by the NIH Toolbox software) by comparing the scores of a test-taker to those in the NIH Toolbox nationally representative normative sample, while adjusting for key demographic variables (age, race, sex, and educational attainment) collected during the Toolbox national norming study.

### Assessment of platelet activation

In sample A, activated platelets (positive for the platelet marker [CD41; Integrin α_IIb_] and platelet activation marker [CD62P; P-selectin]) [25] were identified in fasting venous blood (drawn between 7-9am) by flow cytometry. Flow cytometry was performed within an hour of blood draw by an investigator blinded to the MetS status of the patients.

For the measurement of plasma sP-selectin in sample B, we collected fasting venous blood (drawn between 6 and 8 AM) and separated plasma within 1 h of blood collection. We then aliquoted 0.5 ml plasma into eppendorf® tubes and immediately stored at −80 °C until further analysis. sP-selectin was measured using Enzyme-linked immunosorbent assay (ELISA) kits (RayBiotech, Inc. Georgia, USA), according to the manufacturer’s instructions. The plasma samples were diluted 1:50 and sample optical density values were compared to standard curves ranging from 0–30ng/ml. The sensitivity of the assay was 20 pg/ml.

### Statistical Analyses

Due to the small patient sample sizes, statistical analyses were carried out using non-parametric tests. For demographic variables, we have presented medians and percentages (as appropriate). We compared platelet activation levels (flow cytometry) between MetS+ and MetS-using the Mann Whitney test, and plasma sP-selectin, MCCB and NIH Toolbox scores using the median (exact) test. We then evaluated the relationship between platelet activation and cognition (MCCB and NIH Toolbox) using Spearman’s rho correlation. 2-sided alpha level was set at 0.05. Statistical analyses were performed with IBM SPSS, Version 28 (IBM Corp., Armonk, NY, USA).

## Results

Table 1 shows the demographic and education distribution of study samples A and B. In sample A, the median age for MetS+ was 10 years higher than MetS-(48 years vs 38 years) and MetS-had more individuals with some college education or higher. Participants in sample B had a median age of 31 years.

**Table 1.** Age, sex, race and education distribution of study samples A and B respectively

| **Study sample A** |  |  |
| --- | --- | --- |
|  | Patients with MetS (n=6) | Patients without MetS (n=7) |
| Age in years (median, minimum, maximum) | 48 (31, 55) | 38 (27, 57) |
| Sex |  |  |
| Male | 3 | 5 |
| Female | 3 | 2 |
| Race |  |  |
| Asian | 1 | 0 |
| Black | 4 | 5 |
| White | 1 | 2 |
| Education |  |  |
| High school | 1 | 0 |
| Associates degree | 2 | 0 |
| Some college | 2 | 5 |
| College degree or higher | 1 | 2 |
| **Study sample B** |  |  |
|  | Patients with MetS (n=10) | Patients without MetS (n=10) |
| Age in years (median, minimum, maximum) | 31 (19, 49) | 31 (19, 49) |
| Sex |  |  |
| Male | 7 | 7 |
| Female | 3 | 3 |
| Race |  |  |
| Hispanic | 3 | 3 |
| Black | 4 | 4 |
| White | 3 | 3 |
| Education |  |  |
| Did not complete high school | 2 | 4 |
| High school/GED | 2 | 3 |
| Some college | 5 | 2 |
| College degree or higher | 1 | 1 |

Abbreviation: MetS = metabolic syndrome

### Platelet activation in MetS+ and MetS-

In sample A, platelet activation detected by flow cytometry, was higher in MetS+ than MetS- (mean rank 8.60 vs. 3.83, p=0.017) (Fig. 1.). Plasma sP-selectin median levels did not defer between MetS+ and MetS- in sample B (48.24 ng/ml vs. 40.15 ng/ml, p=0.809).

**Fig. 1.**
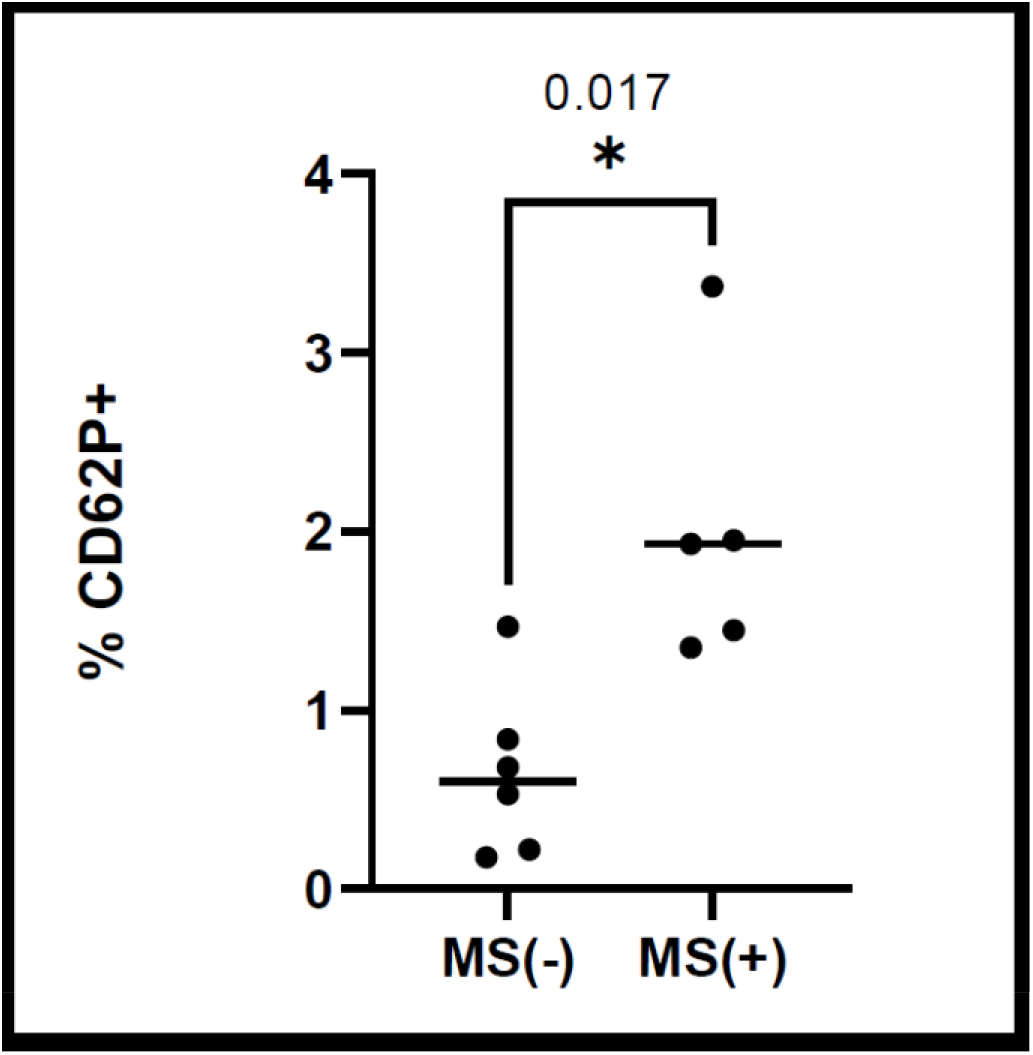
Platelet activation in patients with metabolic syndrome [MS(+)] relative to patients without metabolic syndrome [MS(-)]. Patients with metabolic syndrome, MS(+) have higher levels of activated platelets relative to patients without metabolic syndrome, MS(-). Activated platelets express the CD62P (P-selectin) marker. Blood was drawn between 7 and 9am and analyzed in the flow cytometer within an hour. Blood from 6 patients without metabolic syndrome and 5 with metabolic syndrome were analyzed. *= p-value (0.017).

### Cognitive function scores in MetS+ and MetS-

MCCB individual and composite median scores did not defer between MetS+ and MetS-in sample A (supplementary table 1). However in sample B, picture vocabulary median scores (language abilities) were significantly lower in MetS+ relative to MetS-(82.35 vs. 104, p=0.015) (supplementary table 2).

### Relationship between platelet activation and cognition

In sample A, platelet activation correlated negatively (rho = −0.74, p= 0.009) with the Wechsler Memory Scale: Spatial Span (nonverbal working memory) (Fig. 2 and supplementary table 3). Also, in sample B, sP-selectin correlated negatively (rho = −0.55, p= 0.029) with the List Sorting Working Memory Test (measure of working memory) (Fig. 3 and supplementary table 4).

**Fig. 2.**
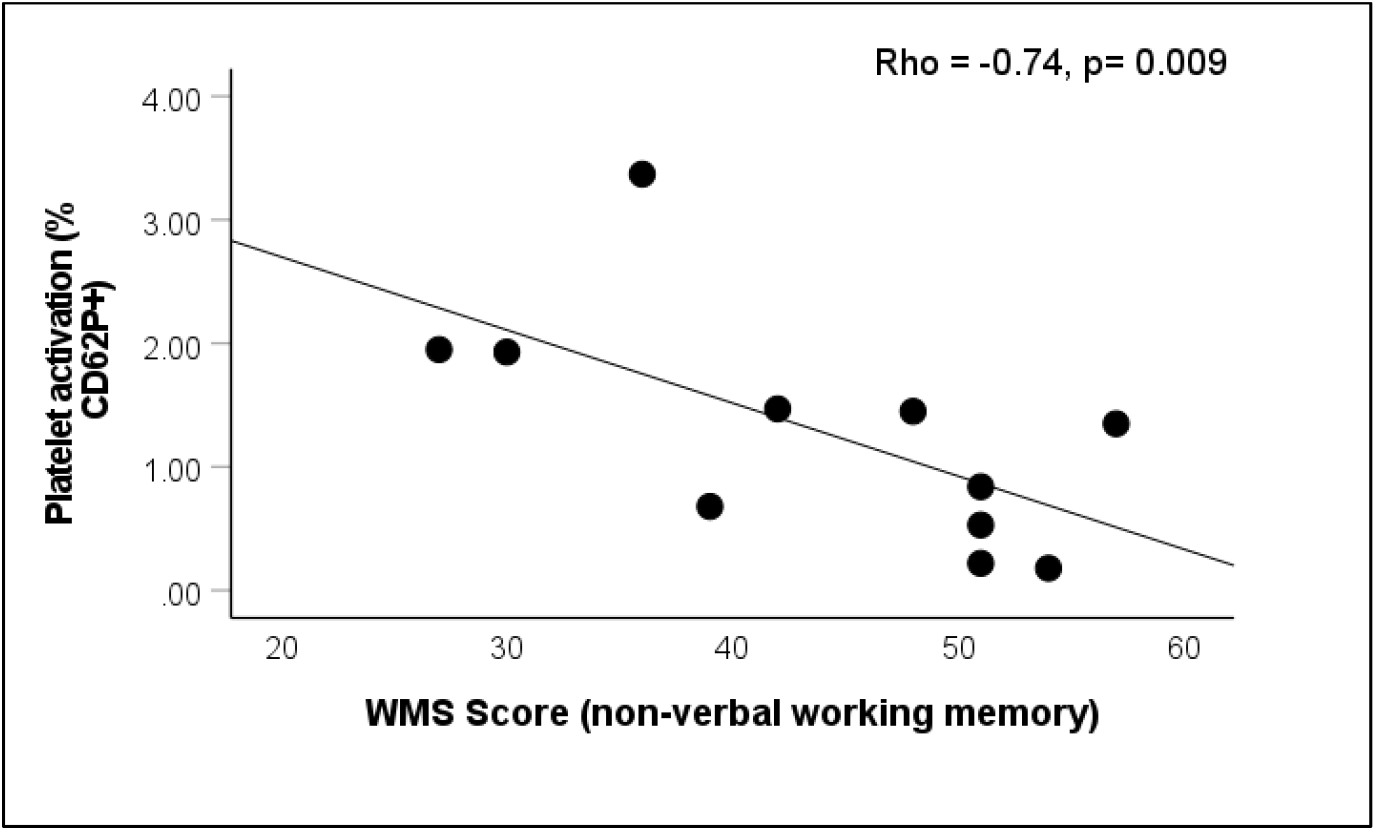
Correlation between platelet activation level and scores on the Measurement and Treatment Research to Improve Cognition in Schizophrenia (MATRICS) Consensus Cognitive Battery (MCCB) Wechsler Memory Scale®—3rd Ed: Spatial Span (WMS). Platelet activation (assessed by flow cytometric detection of platelet activation marker [CD62P; P-selectin]) correlated negatively with scores on the WMS. The WMS is a test of nonverbal working memory. Higher scores on the WMS indicate superior cognition.

**Fig. 3.**
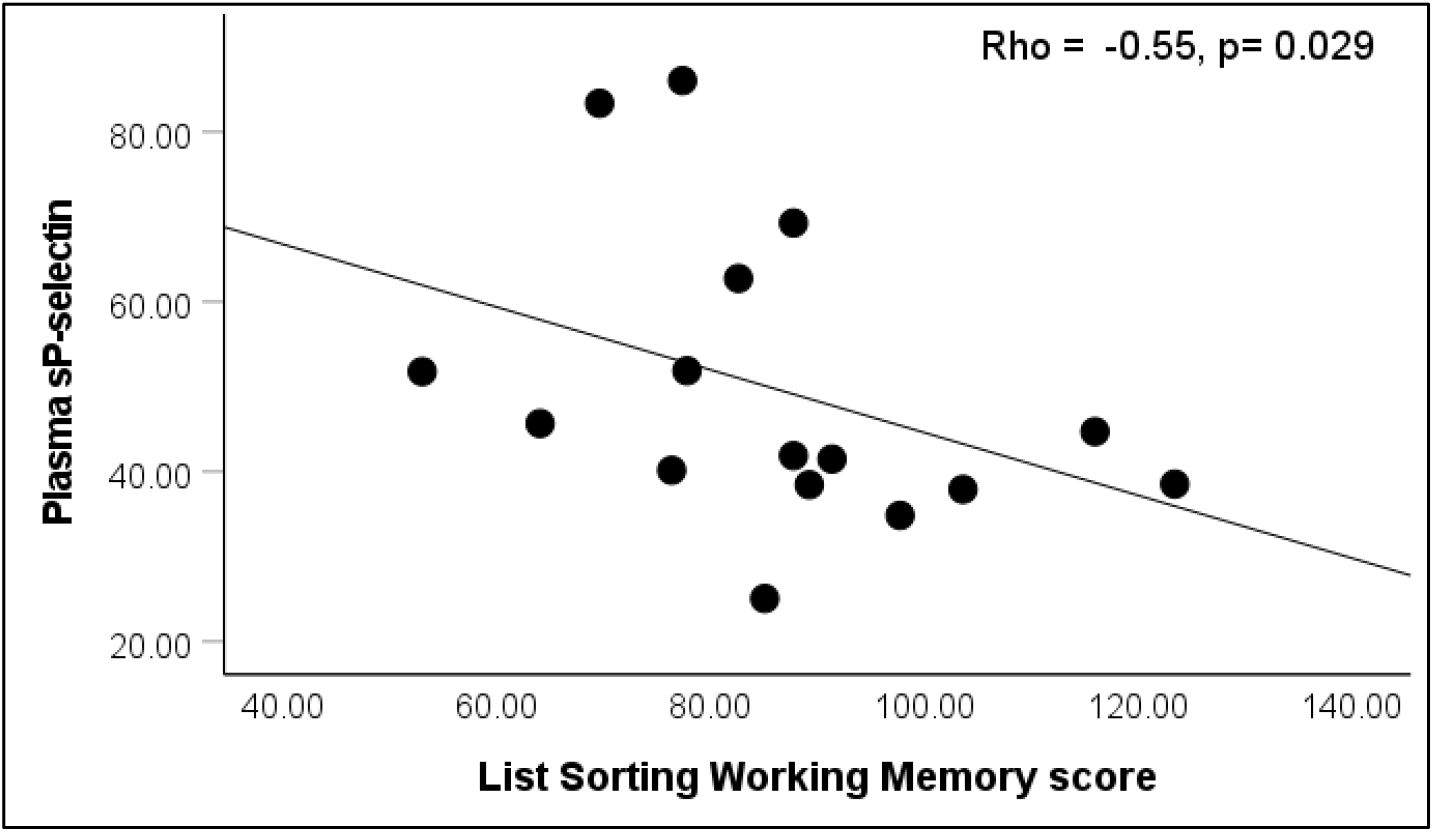
Correlation between plasma sP-selectin and the NIH Toolbox List Sorting Working Memory Test. Plasma sP-selectin negatively correlated with the List Sorting Working Memory test which measures working memory. Higher scores on the List Sorting Working Memory Test indicate superior cognition. Four participants had missing data (i.e. n=16 for the correlational analysis).

## Discussion/Conclusion

We demonstrated the feasibility of flow cytometer-based platelet analysis concurrently with cognitive assessments in patients with schizophrenia and found platelet activation to be associated with both MetS and cognitive impairment in patients with schizophrenia. Specifically, we found higher levels of platelet activation and lower language skills in patients with schizophrenia comorbid with MetS relative to patients without MetS, and a negative correlation between platelet activation and working memory. To our knowledge, our study is the first to concurrently associate platelet activation with MetS and cognitive impairment in patients with schizophrenia.

In contrast to flow cytometry-detected platelet activation, platelet activation assessed by plasma sP-selectin did not differ between MetS+ and MetS-. Although MetS+ had higher median sP-selectin levels, it was not statistically different from MetS-probably due to the small sample size but it could also be a reflection of lower sensitivity of sP-selectin as a marker of platelet activation compared to flow cytometry which directly measures P-selectin on platelets [26]. Interestingly, there was a general pattern of negative correlation between platelet activation assessed either by flow cytometry or sP-selectin and the several cognitive function tests in the MCCB or the NIH Toolbox (supplementary tables 3 and 4). Additionally, the lack of statistical significance in the correlations between platelet activation and the several cognitive tests might be due to small sample size. The significant correlations between platelet activation and working memory in the MCCB and the NIH Toolbox respectively, require replication in larger studies. Our finding of lower language functioning in MetS+ is consistent with findings in non-psychiatric samples [27,28] but, to our knowledge, language function has not been previously reported in patients with schizophrenia comorbid with MetS.

Our preliminary findings are consistent with the notion that platelet activation may be involved in the association of MetS with cognitive impairment believed to contribute to premature dementia in patients with schizophrenia. Indeed, it is plausible to consider platelet activation as a link between MetS and cognitive impairment in patients with schizophrenia considering the several potential mechanisms by which activated platelets can impact the brain [15]. For example, platelets carry several neurotransmitters and other proteins (e.g. reelin) important for neuronal function [14]. Platelets are also involved in neuroplasticity [14]. Importantly, activated platelets are capable of affecting the structural integrity of the BBB [29], penetrate and interact with the cellular components of the BBB matrix including astrocytes [30]. Relatedly, we found a positive correlation between plasma sP-selectin, and the astrocyte marker, S100B in patients with schizophrenia [17]. Moreover, schizophrenia is now considered a neurodevelopmental and neurodegenerative disorder [7] and astrocytes have been shown to contribute to progression of neurodegeneration in neurodegenerative disorders [31].

The strengths of our pilot study include well-characterized participants with diagnoses confirmed with a validated structured interview, and the use of established cognitive test batteries for comprehensive assessment of cognition. Limitations of our pilot study include the cross-sectional design, small sample size, inability to control for potential confounders (age, sex, education, and psychotic symptom severity) and lack of brain imaging data.

In conclusion, our findings demonstrate an association between platelet activation and both MetS and cognitive impairment in patients with schizophrenia. Future studies are needed to elucidate the role of platelets in MetS-related cognitive impairment in patients with schizophrenia. This line of research could result in novel platelet-based (e.g. P-selectin blocking agents [32]) interventions for the prevention and treatment of MetS-related cognitive impairment.

## Data Availability

The data that support the findings of this study are not publicly available, but a minimal data set required to replicate the study findings reported in the article will be made available on request to the corresponding author.

## Acknowledgement

We thank Anh Bui-Thanh for performing the flow cytometry studies and Cristin Rodriguez for the MCCB assessments. We also thank Professor Consuelo Walss-Bass for supervising the measurement of plasma sP-selectin.

## Statement of Ethics

Pilot study A was reviewed and approved by the Institutional Review Board of Baylor College of Medicine (approval # H-48718) and the Research and Development Committee of the Michael E. DeBakey VA Medical Center. Pilot study B was reviewed and approved by The institutional review board of The University of Texas Health Science Center, Houston. Written informed consent was obtained from all the participants prior to participating in both pilot studies.

## Author Contributions

The work presented here was carried out in collaboration between all the authors. OOO designed the study, and wrote the first draft of the manuscript. RER and KVV supervised the flow cytometry experiments and provided critical intellectual input in the several iterations of the manuscript. All authors read and approved the final manuscript.

## References

1 Rs KK, Rs KK, Kahn RS, Keefe RSE. Schizophrenia is a cognitive illness: time for a change in focus. JAMA psychiatry. 2013;70(10):1107–12.

2 Miniawi S El, Orgeta V, Stafford J. Non-affective psychotic disorders and risk of dementia: a systematic review and meta-analysis. Psychol Med. 2022 Oct;1–13.

3 Stroup TS, Olfson M, Huang C, Wall MM, Goldberg T, Devanand DP, et al. Age-Specific Prevalence and Incidence of Dementia Diagnoses Among Older US Adults With Schizophrenia. JAMA psychiatry. 2021 Jun;78(6):632–41.

4 Lang X, Liu Q, Fang H, Zhou Y, Forster MT, Li Z, et al. The prevalence and clinical correlates of metabolic syndrome and cardiometabolic alterations in 430 drug-naive patients in their first episode of schizophrenia. Psychopharmacology (Berl). 2021 Dec;238(12):3643–52.

5 Vancampfort D, Stubbs B, Mitchell AJ, De Hert M, Wampers M, Ward PB, et al. Risk of metabolic syndrome and its components in people with schizophrenia and related psychotic disorders, bipolar disorder and major depressive disorder: a systematic review and meta-analysis. World Psychiatry. 2015 Oct;14(3):339–47.

6 Hagi K, Nosaka T, Dickinson D, Lindenmayer JP, Lee J, Friedman J, et al. Association Between Cardiovascular Risk Factors and Cognitive Impairment in People With Schizophrenia: A Systematic Review and Meta-analysis. JAMA psychiatry. 2021 May;78(5):510–8.

7 Jonas K, Abi-Dargham A, Kotov R. Two Hypotheses on the High Incidence of Dementia in Psychotic Disorders. JAMA psychiatry. 2021 Dec;78(12):1305–6.

8 Marques P, Collado A, Martinez-Hervás S, Domingo E, Benito E, Piqueras L, et al. Systemic Inflammation in Metabolic Syndrome: Increased Platelet and Leukocyte Activation, and Key Role of CX3CL1/CX3CR1 and CCL2/CCR2 Axes in Arterial Platelet-Proinflammatory Monocyte Adhesion. J Clin Med. 2019 May;8(5). DOI: 10.3390/JCM8050708

9 Santilli F, Vazzana N, Liani R, Guagnano MT, Daví G. Platelet activation in obesity and metabolic syndrome. Obes Rev. 2012 Jan;13(1):27–42.

10 Lam FW, Vijayan KV, Rumbaut RE. Platelets and Their Interactions with Other Immune Cells. Compr Physiol. 2015 Jul;5(3):1265–80.

11 Dole VS, Bergmeier W, Patten IS, Hirahashi J, Mayadas TN, Wagner DD. PSGL-1 regulates platelet P-selectin-mediated endothelial activation and shedding of P-selectin from activated platelets. Thromb Haemost. 2007 Oct;98(4):806–12.

12 Kisucka J, Chauhan AK, Zhao B-QQ, Patten IS, Yesilaltay A, Krieger M, et al. Elevated levels of soluble P-selectin in mice alter blood-brain barrier function, exacerbate stroke, and promote atherosclerosis. Blood. 2009 Jun;113(23):6015–22.

13 Wang Q, Zhao W, Bai S. Association between plasma soluble P-selectin elements and progressive ischemic stroke. Exp Ther Med. 2013 May;5(5):1427–33.

14 Burnouf T, Walker TL. The multifaceted role of platelets in mediating brain function. Blood. 2022 Aug;140(8):815–27.

15 Arnoldussen IAC, Witkamp RF. Effects of Nutrients on Platelet Function: A Modifiable Link between Metabolic Syndrome and Neurodegeneration? Biomolecules. 2021 Oct;11(10). DOI: 10.3390/BIOM11101455

16 Walsh MTH, Ryan M, Hillmann A, Condren R, Kenny D, Dinan T, et al. Elevated expression of integrin αIIb βIIIa in drug-naïve, first-episode schizophrenic patients. Biol Psychiatry. 2002 Nov;52(9):874–9.

17 Pinjari OF, Dasgupta SK, Okusaga OO. Plasma Soluble P-selectin, Interleukin-6 and S100B Protein in Patients with Schizophrenia: a Pilot Study. Psychiatr Q. 2022 Mar;93(1):335–45.

18 Rezaianzadeh A, Namayandeh S-M, Sadr S-M. National Cholesterol Education Program Adult Treatment Panel III Versus International Diabetic Federation Definition of Metabolic Syndrome, Which One is Associated with Diabetes Mellitus and Coronary Artery Disease? Int J Prev Med. 2012 Aug [cited 2018 Jul 20].; 3(8):552–8.

19 Hartman LI, Heinrichs RW, Mashhadi F. The continuing story of schizophrenia and schizoaffective disorder: One condition or two? Schizophr Res Cogn. 2019 Jun;16:36.

20 Sheehan D V., Lecrubier Y, Sheehan KH, Amorim P, Janavs J, Weiller E, et al. The Mini-International Neuropsychiatric Interview (M.I.N.I.): The development and validation of a structured diagnostic psychiatric interview for DSM-IV and ICD-10. Journal of Clinical Psychiatry. 1998; pp 22–33.

21 Buchanan RW, Keefe RSE, Umbricht D, Green MF, Laughren T, Marder SR. The FDA-NIMH-MATRICS guidelines for clinical trial design of cognitive-enhancing drugs: what do we know 5 years later? Schizophr Bull. 2011 Nov;37(6):1209–17.

22 Zelazo PD, Anderson JE, Richler J, Wallner-Allen K, Beaumont JL, Conway KP, et al. NIH Toolbox Cognition Battery (CB): validation of executive function measures in adults. J Int Neuropsychol Soc. 2014 Jul;20(6):620–9.

23 Akshoomoff N, Newman E, Thompson WK, McCabe C, Bloss CS, Chang L, et al. The NIH Toolbox Cognition Battery: results from a large normative developmental sample (PING). Neuropsychology. 2014 Jan;28(1):1–10.

24 Heaton RK, Akshoomoff N, Tulsky D, Mungas D, Weintraub S, Dikmen S, et al. Reliability and validity of composite scores from the NIH Toolbox Cognition Battery in adults. J Int Neuropsychol Soc. 2014;20(6):588–98.

25 Khatlani T, Pradhan S, Da Q, Gushiken FC, Bergeron AL, Langlois KW, et al. The β isoform of the catalytic subunit of protein phosphatase 2B restrains platelet function by suppressing outside-in αIIbβ3 integrin signaling. J Thromb Haemost. 2014 Dec;12(12):2089.

26 Morel A, Rywaniak J, Bijak M, Miller E, Niwald M, Saluk J. Flow cytometric analysis reveals the high levels of platelet activation parameters in circulation of multiple sclerosis patients. Mol Cell Biochem. 2017 Jun;430(1–2):69–80.

27 Cahana-Amitay D, Spiro A, Cohen JA, Oveis AC, Ojo EA, Sayers JT, et al. Effects of metabolic syndrome on language functions in aging. J Int Neuropsychol Soc. 2015 Mar;21(2):116–25.

28 Wu SE, Chen WL. Longitudinal trajectories of metabolic syndrome on different neurocognitive domains: a cohort study from the Taiwan biobank. Aging (Albany NY). 2021 Jun;13(11):15400–12.

29 Jones LD, Jackson JW, Maggirwar SB. Modeling HIV-1 Induced Neuroinflammation in Mice: Role of Platelets in Mediating Blood-Brain Barrier Dysfunction. PLoS One. 2016 Mar;11(3). DOI: 10.1371/JOURNAL.PONE.0151702

30 Kniewallner KM, de Sousa DMB, Unger MS, Mrowetz H, Aigner L. Platelets in Amyloidogenic Mice Are Activated and Invade the Brain. Front Neurosci. 2020 Mar;14. DOI: 10.3389/FNINS.2020.00129

31 Brandebura AN, Paumier A, Onur TS, Allen NJ. Astrocyte contribution to dysfunction, risk and progression in neurodegenerative disorders. Nat Rev Neurosci. 2022 DOI: 10.1038/S41583-022-00641-1

32 Karki NR, Kutlar A. P-Selectin Blockade in the Treatment of Painful Vaso-Occlusive Crises in Sickle Cell Disease: A Spotlight on Crizanlizumab. J Pain Res. 2021;14:849– 56.

