## Supplementary Table for "Is platelet activation a link between metabolic syndrome and cognitive impairment in patients with schizophrenia?"

| **MCCB Cognitive Test (median score)** | **MetS+ (n=6)** | **MetS- (n=7)** | **P-value** |
| --- | --- | --- | --- |
| Trail Making Test: Part A (TMT) | 30.50 | 34.00 | 0.592 |
| Brief Assessment of Cognition in Schizophrenia: Symbol-Coding (BACS:SC) | 25.50 | 32.00 | 0.559 |
| Hopkins Verbal Learning Test—Revised™ (HVLT-R) | 29.50 | 30.00 | 0.797 |
| Wechsler Memory Scale®—3rd Ed: Spatial Span (WMS) | 33.00 | 51.00 | 0.592 |
| Letter–Number Span (LNS) | 25.50 | 35.00 | 0.590 |
| Neuropsychological Assessment Battery®: Mazes (NAB:MAZES) | 38.50 | 43.00 | 0.592 |
| Brief Visuospatial Memory Test—Revised (BVMT-R) | 33.00 | 43.00 | 0.592 |
| Category Fluency: Animal Naming | 37.00 | 42.00 | 0.790 |
| Mayer-Salovey-Caruso Emotional Intelligence Test™: Managing Emotions (MSCEIT) | 54.50 | 75.00 | 0.103 |
| Continuous Performance Test—Identical Pairs (CPT-IP) | 28.00 | 34.00 | 0.592 |
| Neurocognitive composite | 24.50 | 32.00 | 0.592 |
| MCCB composite | 24.50 | 37.00 | 0.559 |

Supplementary Table 1: Median MCCB test scores for MetS+ and MetS- in study sample A

Abbreviation: MetS+ = patients with schizophrenia comorbid with metabolic syndrome. MetS- = patients with schizophrenia without metabolic syndrome. MCCB = the Measurement and Treatment Research to Improve Cognition in Schizophrenia (MATRICS) Consensus Cognitive Battery. MetS was based on the National Cholesterol Education Program Adult Treatment Panel III (NCEP ATP III) criteria.

Supplementary Table 2. Median NIH Toolbox cognitive test scores for MetS+ and MetS- in study sample B

| **NIH Toolbox Cognitive Test (median score)** | **MetS+ (n=10)** | **MetS- (n=10)** | **P-value** |
| --- | --- | --- | --- |
| Picture Sequence Memory Test | 68.28 | 75.27 | 0.153 |
| Pattern Comparison Processing Speed | 70.86 | 68.81 | 0.772 |
| Dimensional Change Card Sort | 81.56 | 86.77 | 0.614 |
| List Sorting Working Memory Test | 86.43 | 85.08 | 0.772 |
| Flanker Inhibitory Control and Attention Test | 98.71 | 78.160 | 0.335 |
| Picture Vocabulary Test | 82.35 | 104.00 | **0.015** |
| Oral Reading Recognition Test | 122.10 | 113.83 | 0.335 |
| Cognition Crystallized Composite | 103.10 | 97.10 | 0.608 |
| Cognition Fluid Composite | 71.59 | 60.09 | 0.614 |
| Cognition Total Composite Score | 87.87 | 69.72 | 0.608 |

Supplementary Table 3. Correlations between platelet activation (flow cytometry) and the individual MCCB test scores (T-scores) in study sample A

|  |  | TMT | BACS:SC | HVLT-R | WMS | LNS | NAB:  MAZES | BVMT-R | Fluency | MSCEIT | CPT-IP | Neuro  cognitive composite score | MCCB composite score |
| --- | --- | --- | --- | --- | --- | --- | --- | --- | --- | --- | --- | --- | --- |
| Platelet activation | Rho | -0.39 | -0.29 | 0.28 | **-0.74** | -0.07 | -0.59 | -0.17 | 0.33 | -0.40 | -0.44 | -0.24 | -0.31 |
|  | p-value | 0.239 | 0.383 | 0.406 | **0.009** | 0.831 | 0.057 | 0.611 | 0.326 | 0.230 | 0.175 | 0.483 | 0.361 |

Abbreviation: TMT = Trail Making Test: Part A; BACS:SC = Brief Assessment of Cognition in Schizophrenia: Symbol-Coding; HVLT-R = Hopkins Verbal Learning Test—Revised™; WMS = Wechsler Memory Scale®—3rd Ed: Spatial Span; LNS = Letter–Number Span; NAB:MAZES = Neuropsychological Assessment Battery®: Mazes; BVMT-R = Brief Visuospatial Memory Test—Revised; Fluency = Category Fluency: Animal Naming; MSCEIT = Mayer-Salovey-Caruso Emotional Intelligence Test™: Managing Emotions; CPT-IP = Continuous Performance Test—Identical Pairs.

Supplementary Table 4: Correlation between plasma soluble P-selectin and the individual NIH Toolbox cognitive measures in study sample B.

|  |  | OR | PS | PC | DC | LS | FL | PV | CC | FC | TC |
| --- | --- | --- | --- | --- | --- | --- | --- | --- | --- | --- | --- |
| Plasma  sP-selectin | Rho | -0.25 | 0.06 | -0.10 | -0.20 | **-0.55** | 0.39 | 0.01 | -0.13 | -0.01 | 0.13 |
|  | p-value | 0.339 | 0.837 | 0.712 | 0.483 | **0.029** | 0.141 | 0.974 | 0.657 | 0.960 | 0.648 |

Abbreviation: OR = Oral Reading Recognition Test; PS = Picture Sequence Memory Test; PC = Pattern Comparison Processing Speed Test; DC = Dimensional Change Card Sort Test; LS = List Sorting Working Memory Test; FL = Flanker Inhibitory Control and Attention Test; PV = Picture Vocabulary Test; CC = Crystallized Cognition Composite Score; FC = Fluid Cognition Composite Score; TC = Total Cognition Composite Score.
